# A Comparative Performance Analysis of Regular Expressions and an LLM-Based Approach to Extract the BI-RADS Score from Radiological Reports

**DOI:** 10.1101/2025.06.01.25328636

**Authors:** Fabio Dennstädt, Luc Lerch, Max Schmerder, Nikola Cihoric, Grazia Maria Cereghetti, Roberto Gaio, Harald Bonel, Irina Filchenko, Janna Hastings, Florian Dammann, Daniel M Aebersold, Hendrik von Tengg-Kobligk, Knud Nairz

## Abstract

**Background:** Different Natural Language Processing (NLP) techniques have demonstrated promising results for data extraction from radiological reports. Both traditional rule-based methods like regular expressions (Regex) and modern Large Language Models (LLMs) can extract structured information. However, comparison between these approaches for extraction of specific radiological data elements has not been widely conducted.

**Methods:** We compared accuracy and processing time between Regex and LLM-based approaches for extracting BI-RADS scores from 7,764 radiology reports (mammography, ultrasound, MRI, and biopsy). We developed a rule-based algorithm using Regex patterns and implemented an LLM-based extraction using the Rombos-LLM-V2.6-Qwen-14b model. A ground truth dataset of 199 manually classified reports was used for evaluation.

**Results:** There was no statistically significant difference in the accuracy in extracting BI-RADS scores between Regex and an LLM-based method (accuracy of 89.20% for Regex versus 87.69% for the LLM-based method; p=0.56). Compared to the LLM-based method, Regex processing was more efficient, completing the task 28,120 times faster (0.06 seconds vs. 1687.20 seconds). Further analysis revealed LLMs favored common classifications (particularly BI-RADS value of 2) while Regex more frequently returned “unclear” values. We also could confirm in our sample an already known laterality bias for breast cancer (BI-RADS 6) and detected a slight laterality skew for suspected breast cancer (BI-RADS 5) as well.

**Conclusion:** For structured, standardized data like BI-RADS, traditional NLP techniques seem to be superior, though future work should explore hybrid approaches combining Regex precision for standardized elements with LLM contextual understanding for more complex information extraction tasks.

## INTRODUCTION

### Natural Language Processing for Data Extraction from Radiology Reports

The use of Natural Language Processing (NLP) has been of long-standing interest for extraction of data from medical reports, particularly in radiology. Traditional rule-based NLP techniques such as regular expressions (Regex) have shown good success in extracting structured data from radiological documents. As for example, Sippo et al. used a Regex-based approach to extract BI-RADS from different radiology reports and achieved a recall of 100.0% and a precision of 96.6% for correct identification of BI-RADS final assessment categories [1].

Recently, large language models (LLMs) as a form of generative artificial intelligence (AI) are increasingly being used for data extraction from radiological reports. Instead of relying on hard-coded rules, LLMs produce token sequences based on given input, which can be used to answer questions about a given text document and thereby extract relevant information. LLMs opened new opportunities for such tasks that traditionally require human-level reasoning and context understanding. LLMs have been shown to be useful in structuring radiology reports [2] as well as for extraction of data from various documents including brain MRI reports [3], chest x-rays and chest CT reports [4], and sonography reports [5]. Overall, LLMs are increasingly implemented in healthcare [6] and used for data extraction in radiology [7].

As we had seen in one of our own recent studies, using locally deployed LLMs within a resource-light, easy-to-use framework, can achieve high levels of performance in extracting various relevant information from mammography reports [8]. However, even when using less resource-demanding LLMs, these models still require considerable hardware infrastructure and computational resources compared to traditional NLP techniques.

Both Regex and LLM-based data extraction have been applied to extraction of BI-RADS from radiological reports, representing critical information that can be retrieved with both systems. However, to our knowledge there has not yet been a direct comparison in a scientific study. In this work we therefore compared the effectiveness of a Regex method versus an LLM-based approach for extracting BI-RADS scores from different radiological reports.

## METHODS

### Dataset

A dataset of 7,764 anonymized, free-text radiology reports from two Swiss hospitals were used in our study. All reports were in German language. The dataset included 5,358 mammography, 1,153 breast ultrasound, 1,150 breast MRI, and 100 radiological biopsies procedure reports.

To obtain a ground truth dataset for the evaluation, the BI-RADS scores were manually classified for a subset of 199 randomly selected reports. Two researchers independently assigned the values for the reports with the possible values “0-6” or “unclear” for each side. The value “unclear” was assigned for cases where the BI-RADS score was not mentioned in the report, or the correct value for a side was not clear. A differentiation between these options was not done, since realization with Regex appears very complex and of limited additional value. In cases of disagreement between the two researchers doing the manual classification a final decision was made by the study coordinators.

### Regex

A rule-based algorithm utilizing Regex to extract BI-RADS scores was developed. The algorithm targeted variations in how BI-RADS scores and side indicators (left, right, or both) were described in the reports. The Regex pattern used to identify BI-RADS scores accounted for numeric scores (0–6) with optional suffixes (e.g., “4a”), as well as Roman numeral scores (I–VI) with optional suffixes. Variants of the term “BI-RADS” (e.g., “BIRADS,” or “bi-rads” were included. Additionally, proximity-based matching of laterality indicators (“left,” “right,” or “both”) to BI-RADS scores was considered. A Python script for assigning a given score to its laterality based on the Regex patterns was created. The script for the full Regex patterns and extraction logic is provided online [9].

### LLM-based approach

LLMs were used for extraction of BI-RADS in analogy to the approach reported in our previous work [8]. We used the *general-classifier* Python package [10] (version 0.1.10), which is an open-source library for LLM-based text classification based on user-defined topics and for a user-defined model. Calculating the probabilities of each token sequence associated to one of the categories, the output of the LLM is constrained to classify a text (e.g., a mammography report). By defining appropriate topics (BI-RADS left breast and BI-RADS right breast) with corresponding categories (“0-6” and “unclear”), it can extract BI-RADS scores from a given text.

Since BI-RADS scores were also extracted in our previous study (on another dataset), we used the same approach with an analogous instruction prompt for the model to conduct the extraction. The open-source LLM Rombos-LLM-V2.6-Qwen-14b [11] was used in this study. It was selected since it previously had the best performance compared to four other LLMs deployable on a single GPU (accuracy of 95.08% in extracting BI-RADS scores from mammography reports) [8]. Rombos-LLM-V2.6-Qwen-14b is a fine-tuned version of Alibaba’s Qwen2.5-14B model for precise instruction following with 14.8 billion parameters and a size of 29.57 GB. A Python script was created to conduct the LLM-based data extraction, which is available on GitHub [9].

Both approaches (Regex and LLM-based) were applied to both the ground truth dataset as well as the entire dataset. The Python scripts were executed on a local Linux server with an RTX6000 Nvidia GPU (with 48GB RAM).

### Statistical Analysis

The Inter-rater agreement (IRA) between the two researchers doing the manual classification was calculated using Cohen’s Kappa [12]. For comparison of accuracy between Regex and LLM-based approaches, a McNemar Test was used to check for statistically significant difference, while for comparison of individual categories of BI-RADS we used a Fisher’s Exact Test. To adjust for multiple comparisons between the categories, we applied a Bonferroni correction.

## RESULTS

### Ground Truth Dataset and Inter-Rater Agreement

A high level of IRA with an overall Cohen’s Kappa of 0.86 (0.89 for left and 0.80 for right breast) was observed between the two researchers doing the manual classification.

For both sides most reports in the ground-truth dataset were classified as BI-RADS 2 (45.2% left breast; 45.7% right breast) or BI-RADS 3 (25.6% left breast; 22.6% right breast). Classifications as “unclear” were made in 8.5% for the left breast and 9.0% for the right breast. 128 of the reports were mammography reports, 52 sonography reports, 12 MRI reports and 7 biopsy reports.

### Comparison of Accuracy and Time

The regex-based approach achieved an accuracy of 89.20% on the sample subset compared to 87.69% of the LLM-based approach (p=0.56) in extracting the BI-RADS scores of both the left and the right breast. Details of the performance on the different subsets are provided in **Table 1**.

**Table 1:**
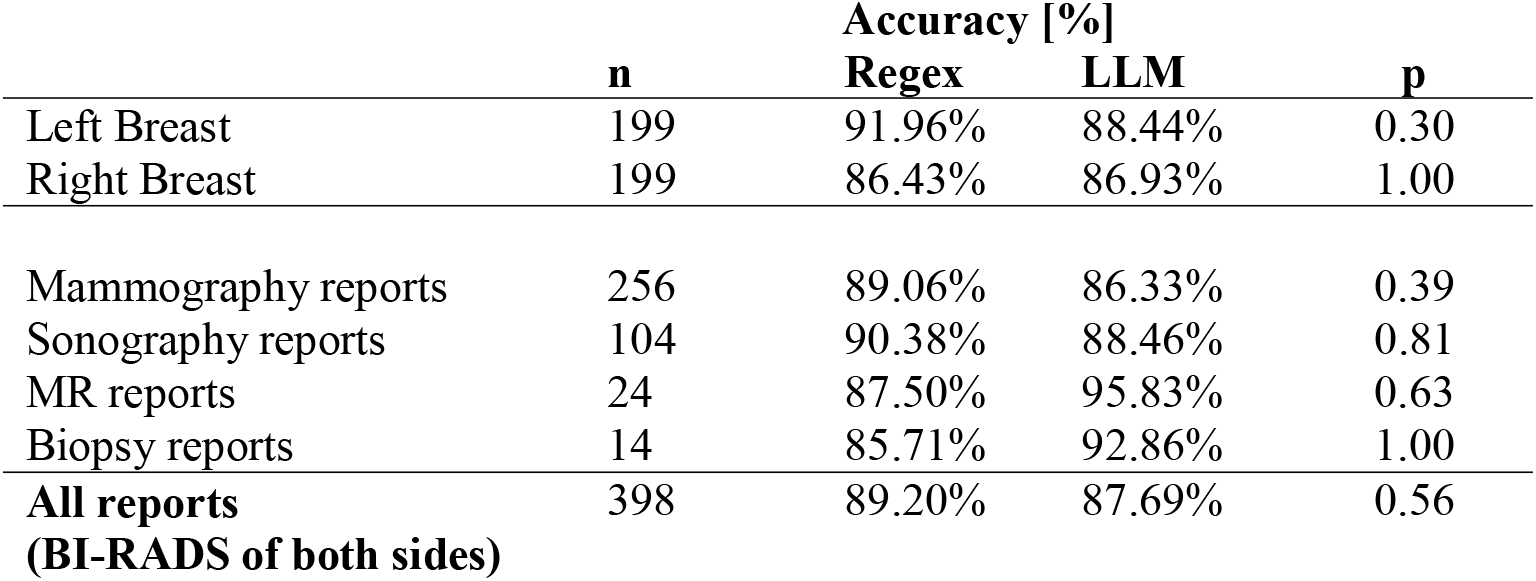
Accuracies of Regex and LLM-based data extraction on different subsets of the data. BI-RADS was extracted two times (left and right side) for each report.

|  |  | Accuracy [%] |  |  |
| --- | --- | --- | --- | --- |
|  | n | Regex | LLM | p |
| Left Breast | 199 | 91.96% | 88.44% | 0.30 |
| Right Breast | 199 | 86.43% | 86.93% | 1.00 |
| Mammography reports | 256 | 89.06% | 86.33% | 0.39 |
| Sonography reports | 104 | 90.38% | 88.46% | 0.81 |
| MR reports | 24 | 87.50% | 95.83% | 0.63 |
| Biopsy reports | 14 | 85.71% | 92.86% | 1.00 |
| **All reports**<br>**(BI-RADS of both sides)** | 398 | 89.20% | 87.69% | 0.56 |

Figure 1. shows the distribution of extracted scores with Regex and LLM compared to the ground truth.

**Figure 1.**
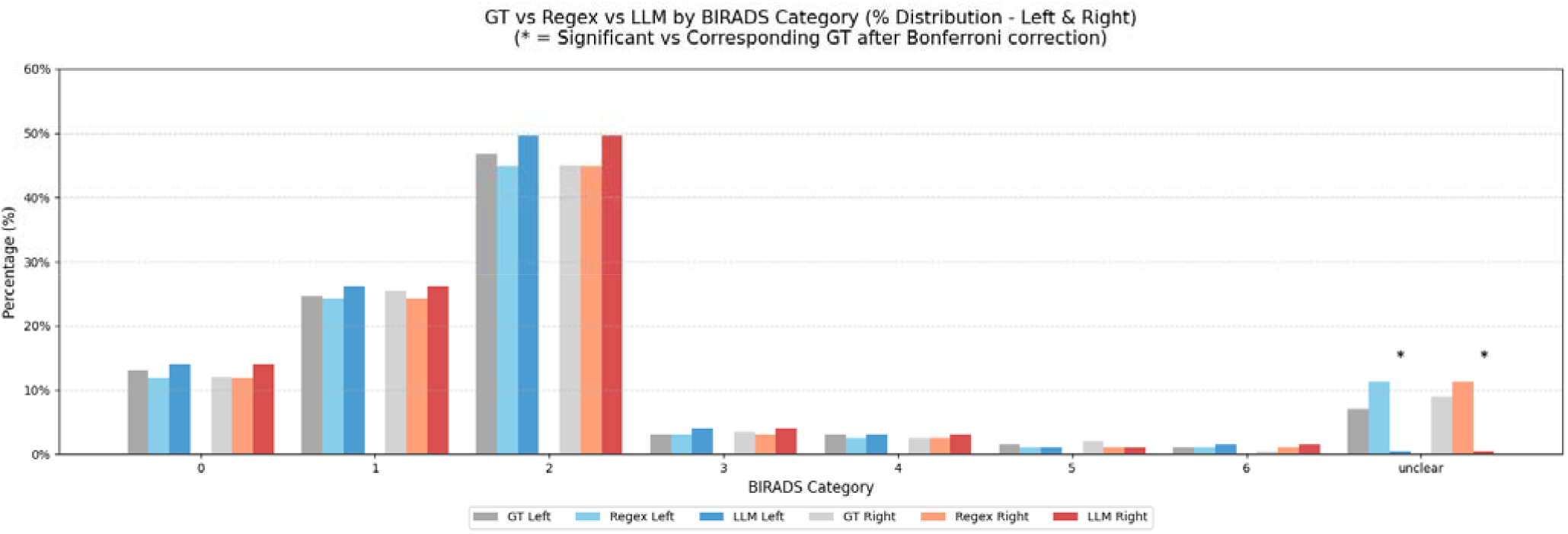
Distribution of scores using Regex and LLM-based data extraction for the left and the right breast on the ground truth dataset. Sub-categories 4a, 4b and 4c were counted as 4. GT: ground truth.

Regarding the required time for conducting the extraction on all reports, the Regex approach required 0.06 seconds compared to 1,687.20 seconds required by the LLM-based approach, meaning the Regex approach was 28,120 times faster to execute.

### Application on Large Clinical Dataset

Similar score distributions as in the ground truth dataset were observed when the extraction methods were applied to the entire datasets of 7,764 radiology reports. A statistically significant difference between left and right side was seen with both approaches for a BI-RADS score of 6 with more instances in the left breast (124 vs. 83; p=0.04 with Regex and 141 vs. 98; p=0.05 with LLM). With the Regex approach a significant difference was also observed for “unclear” BI-RADS scores (901 vs. 755; p<0.01). The results are presented in **Supplementary Figure 1** and **Supplementary Figure 2**.

Significant differences between Regex and LLM-based approach were seen for a BI-RADS score of 0 for the right breast (1066 vs. 1256, p<0.01), BI-RADS score of 2 for both sides (left 3588 vs. 4058, p<0.01; right 3604 vs. 4087, p<0.01), BI-RADS score of 4 for the right breast (256 vs. 325) and “unclear” for both sides (left 755 vs. 87, p<0.01; right 901 vs. 102, p<0.01). The comparison between Regex and LLM-based approach is presented in **Figure 2**. The Regex approach required 1.45 seconds compared to 26,686.44 seconds required by the LLM-based approach, meaning the Regex approach was 18,404.44 times faster.

**Figure 2.**
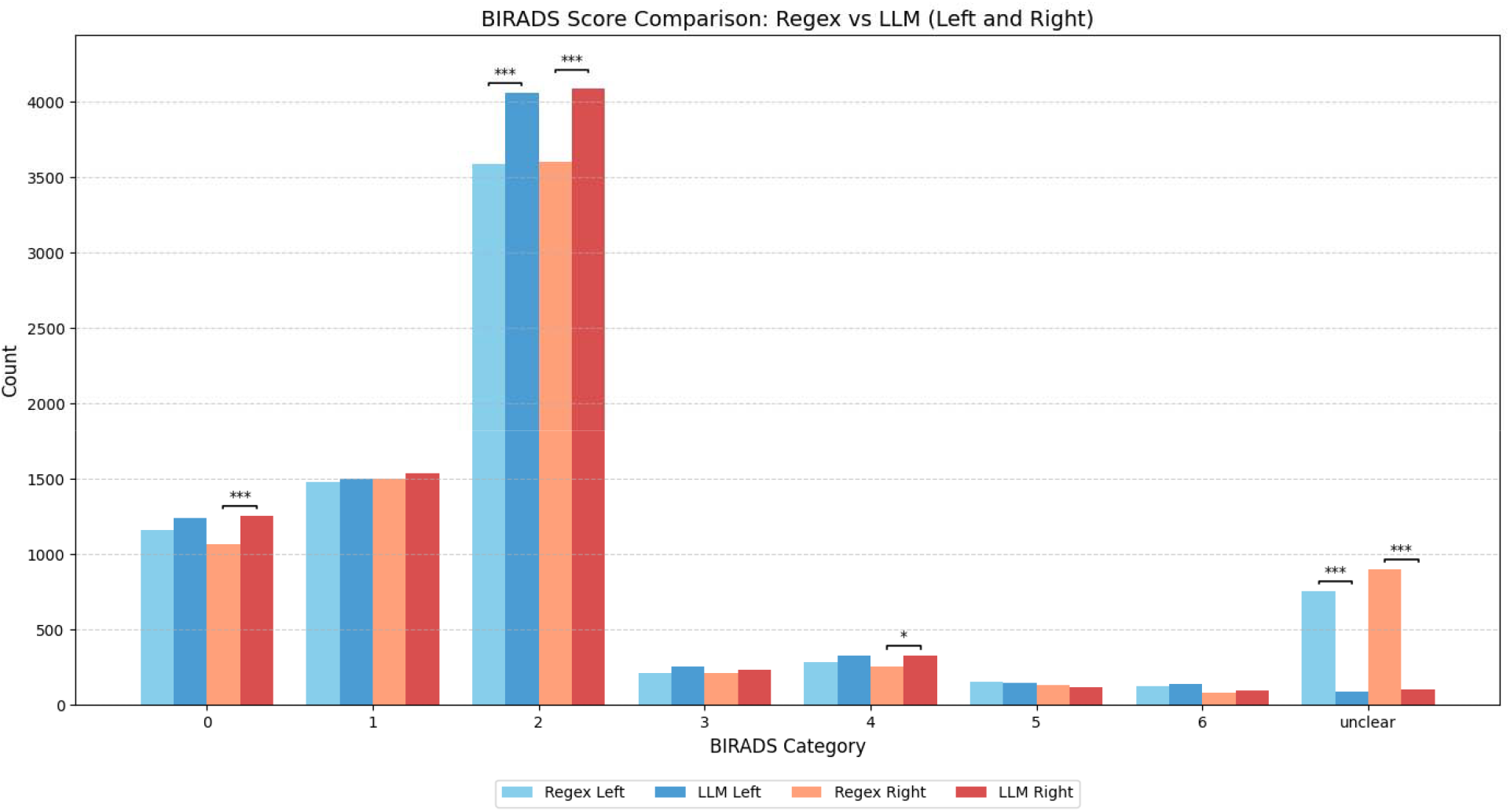
Distribution of scores using Regex and LLM-based data extraction for the left and the right breast on the entire clinical dataset. Sub-categories 4a, 4b and 4c were counted as 4.

## DISCUSSION

### Characteristics, Strengths and Issues of Regex and LLM-based Data Extraction

Our study showed similar accuracies between Regex and LLM-based extraction of BI-RADS scores from different radiological reports. However, Regex were many times faster than LLM-based extraction. Regex is a straightforward approach for extracting clearly defined information from free text. The method has been successfully used for data extraction in radiology for a long time [1], [13]. However, the application for more complex data extraction tasks is limited, as information in free text is often not provided in a defined structured or semi-structured way. BI-RADS is a standardized score, which should be provided clearly written within a radiological report investigating breast cancer [14], [15]. This standardization makes BI-RADS scores well-suited for Regex-based extraction methods. We were able to successfully apply both extraction methods to our heterogeneous dataset of mammography, sonography, MRI and biopsy reports, since standardized terms and scores were used.

For more complex data provided not is such a structured way in a radiology report (e.g., morphological description of lesions or calcifications), context-understand and reasoning are required to accurately extract information. With the technological advancements of LLMs, these models are increasingly applied for extraction of data from free-text reports in radiology with promising results [7]. However, for some data elements more traditional approaches are equally accurate and way more efficient. The best approach may therefore be using a hybrid-system and select the extraction method depending on the relevant information and the datasets used.

Both Regex and LLM-based strategies have different strengths and weaknesses in our dataset. The LLM-based approach more often extracted the value “2”, which was the most frequent value extracted overall and less frequently extracted the value “unclear”. The issue of “LLM hallucinations”, meaning that LLMs produce incorrect statements not backed by data, has been widely reported and is a still largely unresolved problem [16]. Potentially the LLM occasionally fabricates information, providing results such as “BI-RADS=2” (the statistically most common classification and therefore probably also most frequent in the training data of the LLM) even when no supporting evidence exists in the text. Conversely, the LLM rarely returns “BI-RADS=unclear” when faced with ambiguous or missing information. This is also a commonly known issue of LLMs as they struggle to “admit” that they cannot answer a question [17].

Regex on the other side is very unlikely to extract a BI-RADS score from a text that is not present (unless the Regex-rule is inappropriate, or the text contains related text parts that could be confused with the true BI-RADS score). However, if the Regex pattern is not matched the system will provide an “unclear” value, which happens therefore way more often than with the LLM-based approach.

Apart from the performance metrices itself, these are important factors to keep in mind, since it means that the two approaches are more likely to make different kinds of systematic errors.

In addition, it should be noted that a custom Regex needs to be created for each data value to be extracted from a clinical note, which may be time-consuming to develop and require technical expertise, while the LLM-based approach only requires a natural language prompt which any clinician can develop themselves.

### Automated Data Extraction for Analysis of Large Clinical Datasets

Analysis of laterality distributions across BI-RADS categories revealed a statistically significant left-sided predominance for BI-RADS 6, suggesting a higher frequency of unclear or highly suspicious findings on the left side. This pattern supports existing literature on the greater incidence of left-sided breast cancer [18], [19], [20].

Using automated classifications and analysis of medical reports is obviously very interesting for large data analyses. For practical application, efficiency and hardware requirements obviously need to be considered. Based on our results for the use-case of extracting the BI-RADS score from several thousand reports it seems that the Regex approach is generally preferable for this structured data extraction task.

### Limitations

While this study offers a detailed comparison between Regex and LLM-based approaches for BI-RADS score extraction, several limitations must be acknowledged. The dataset was sourced from two institutions within a single regional context, which may limit the generalizability of the findings to other clinical environments with different reporting conventions or languages. Interestingly, the accuracy of the LLM-based approach was lower than in our previous study (87.69 % vs. 95.08 %) [21]. This is most likely because the dataset in the current study contained more complex reports from two different institutions. It may also explain, why the performance in our study is lower than in the study of Sippo et al. [1]. In a recent study by Güneş et al., much larger, modern LLMs of private companies (including GPT-4o, Perplexity and Claude 3.5 Sonnet) were used to answer multiple-choice questions related to BI-RADS and achieved a maximum accuracy of 90% [22]. While the results are not directly comparable, an accuracy of 85-95% seems realistic using current LLM systems.

The Regex-based method achieved high accuracy but used complex patterns addressing different possibilities how the score might be written in a report. While it demonstrated efficiency in structured tasks, its patterns do not account for all possible linguistic variations in mammography reports or possible types (e.g., abbreviations like “Lt” vs. “left,” or ambiguous phrasing like “left greater than right”).

Similarly, the LLM prompts were not optimized to maximize contextual accuracy, potentially leading to misinterpretations of nuanced descriptors. Possible optimization techniques might be additional prompt engineering, few-shot learning or fine-tuning of models. In our study no explicit optimization for either approach was conducted.

Furthermore, ground truth annotations were manually created on a limited subset of the data, potentially reducing the breadth of variability captured in the accuracy assessment. Overall, these limitations highlight the value of exploring hybrid approaches that combine the structural precision of regex with the adaptability of fine-tuned LLMs.

## CONCLUSIONS

Despite the impressive capabilities of LLMs in extracting data from radiology reports, a Regex-based approach was much faster without being less accurate in extracting BI-RADS scores in our study. For structured data like BI-RADS, traditional NLP techniques seem to be superior, though future work should explore hybrid approaches combining Regex precision for standardized elements with LLM contextual understanding for more complex information extraction tasks. Addressing real-life challenges (such as limited hardware resources), will also be crucial for clinical implementation of these technologies.

## List of abbreviations

AI: Artificial Intelligence
BI-RADS: Breast Imaging-Reporting and Data System
GT: ground truth
IRA: Inter-rater agreement
LLM: Large Language Model
MRI: Magnet Resonance Imaging
NLP: Natural Language Processing
Regex: Regular Expressions

## Code and data availability statement

The source code for the Regex and LLM-based approach are provided at GitHub [9]. Information regarding the used anonymized mammography reports can be obtained from the authors upon reasonable request.

## Author contributions

Conceptualization – FD, LL, MS, NC, KN, GMC, HVTK, DMA, JH

Methodology, technical implementation – LL, FD, MS, RG, JM, LC

Methodology, creation of datasets – LL, FD, MS, FD, HB

Statistical Analysis – FD, LL, MS, IF

Writing, original draft preparation – FD, LL, MS, NC

Writing, illustrations – FD, IF

Writing, review and editing – All authors

Project administration – FD, DMA, HVTK, KN

## Acknowledgement

Not applicable.

## Funding

This study was supported by Innosuisse grant 59228.1/120.503 “SMARAGD”.

## Disclosure of potential conflicts of interest

Dr. Cihoric is a technical lead for the *SmartOncology* project and medical advisor for Wemedoo AG, Steinhausen AG, Switzerland.

The authors declare no other conflicts of interest.

## Ethics approval

An ethics approval for the study was granted by the Ethics Committee of the Canton of Bern (BASEC number 2022-01621), in alignment with the principles outlined in the Declaration of Helsinki.

## Informed consent

The data utilized in this study comprised anonymized radiology reports. All patients provided informed consent for the use of their data for research purposes, in accordance with ethical standards.

## Notes

### Author Declarations

Ethics Committee of the Canton of Bern gave ethical approval for this work (BASEC number 2022-01621).

