## Supplementary Figures for "A Comparative Performance Analysis of Regular Expressions and an LLM-Based Approach to Extract the BI-RADS Score from Radiological Reports"

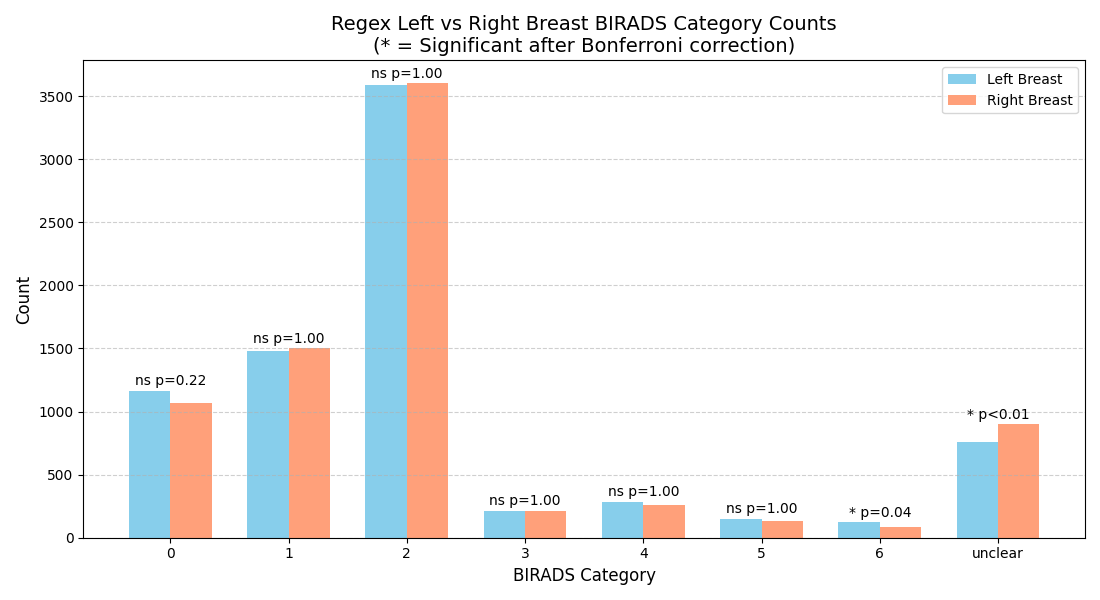

Supplementary Figure 1: Distribution of scores using Regex for data extraction. Sub-categories 4a, 4b and 4c were counted as 4.

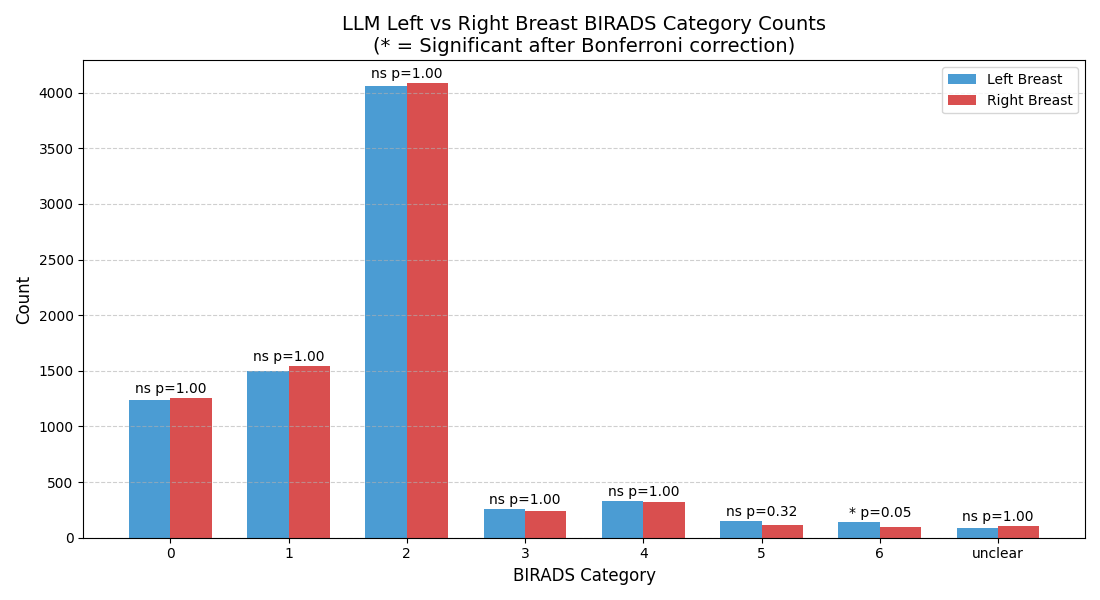

Supplementary Figure 2: Distribution of scores using LLM-based data extraction. Sub-categories 4a, 4b and 4c were counted as 4.
